# Effects of Diabetes and Blood Glucose on COVID-19 Mortality: A Retrospective Observational Study

**DOI:** 10.1101/2021.01.21.20202119

**Authors:** Fei Li, Yue Cai, Chao Gao, Lei Zhou, Renjuan Chen, Kan Zhang, Weiqin Li, Ruining Zhang, Xijing Zhang, Duolao Wang, Yi Liu, Ling Tao

## Abstract

**OBJECTIVE:** To investigate the association of diabetes and blood glucose on mortality of patients with Coronavirus Disease 2019 (COVID-19).

**RESEARCH DESIGN AND METHODS:** This was a retrospective observational study of all patients with COVID-19 admitted to Huo-Shen-Shan Hospital, Wuhan, China. The hospital was built only for treating COVID-19 and opened on February 5, 2020. The primary endpoint was all-cause mortality during hospitalization.

**RESULTS:** Among 2877 hospitalized patients, 13.5% (387/2877) had a history of diabetes and 1.9% (56/2877) died in hospital. After adjustment for confounders, patients with diabetes had a 2-fold increase in the hazard of mortality as compared to patients without diabetes (adjusted HR 2.11, 95%CI: 1.16-3.83, *P*=0.014). The on-admission glucose (per mmol/L≥4mmol/L) was significantly associated with subsequent mortality on COVID-19 (adjusted HR 1.17, 95%CI: 1.10-1.24, *P*<0.001).

**CONCLUSIONS:** Diabetes and on-admission glucose (per mmol/L≥4mmol/L) are associated with increased mortality in patients with COVID-19. These data support that blood glucose should be properly controlled for possibly better survival outcome in patients with COVID-19.

## INTRODUCTION

Coronavirus disease 2019 (COVID-19) is a current pandemic disease caused by the positive-sense RNA virus named severe acute respiratory syndrome coronavirus 2 (SARS-CoV-2). The mortality rates were reported to range from 1% to more than 5% ^1^. Diabetes has been suggested as one of the most common comorbidities and associated with higher mortality. Earlier reports with limited sample sizes, reported that diabetes had a hazard ratio of2-3 for mortality^2,3^. Later on, summarized by the Chinese Center for Disease Control and Prevention, among 72,314 patients with COVID-19 in China, those with diabetes had a mortality of 7.3%compared with that of 2.3% in the general infected population^1^. However, none of these previous studies have adjusted for confounding factors, such as age, hypertension, or cardio-cerebrovascular diseases, which have been previously identified as predictors of COVID-19 related death^2,4^.

On-admission glucose level has been identified to be associated with in-hospital adverse events in patients with pneumonia ^5-7^. Compared with common pneumonia, the outbreak and pandemic of COVID-19 could have much more impact the glucose level in diabetes. As to the patients, anxiety and panic could lead to stress hyperglycemia, and as to the doctor, high workload could result in the inadequate glucose management, and finally as to the SARS-CoV-2, it could impact the glucose metabolism directedly. However, whether the dysglycemia from COVID-19 and the corresponding anti-diabetic treatments have an impact on the mortality in patients with COVID-19 have not been investigated.

Here, we investigated in 2877 patients, consecutively admitted to Huo Shen Shan Hospital, dedicated solely to the treatment of COVID-19, in Wuhan, China. We investigated the association of diabetes, admission glucose and anti-diabetic medications with COVID-19 mortality.

## RESEARCH DESIGN AND METHODS

### Study Participants

This was a retrospective observational study comparing association of both diabetic status and antidiabetic treatment with mortality among hospitalized patients with COVID-19. All patients admitted to Huo Shen Shan Hospital, Wuhan, China, from February 5, 2020, to March 15, 2020, with laboratory-confirmed COVID-19 were included in this study. Huo Shen Shan Hospital was specially built with a capacity of one-thousand beds and opened by the government on February 5th, 2020, dedicated to treating exclusively COVID-19 patients. Patients with COVID-19 included in this study were diagnosed according to World Health Organization interim guidance ^8^.

This study was approved by the National Health Commission of China and the Institutional Review Board at Huo Shen Shan Hospital (Wuhan, China) (HSSLL025). Written informed consent was waived by the Ethics Committee of the Huo Shen Shan Hospital for patients with emerging infectious diseases.

### Data Collection

Patients’ demographic characteristics and clinical data (symptoms, comorbidities, laboratory findings, and outcomes) during hospitalization were collected from electronic medical records by two investigators (R.Z. and Y.C). All data were independently reviewed and entered into the computer database by two analysts (C.G. and Y.C.).

The laboratory diagnoses of COVID-19 were confirmed by the Viral Nucleic Acid Kit ^8^ according to the kit instructions. A 2019-nCoV detection kit (Bioperfectus) was used to detect the ORF1ab gene (nCovORF1ab) and the N gene (nCoV-NP) according to the manufacturer’s instructions, using real-time reverse transcriptase–polymerase chain reaction ^9^. An infection was considered laboratory-confirmed if both the nCovORF1ab and nCoV-NP tests showed positive results.

### Clinical Endpoints

The primary endpoint was all-cause mortality during hospitalization. Secondary endpoints included the use of invasive mechanical ventilation, and the severity of the COVID-19. The severity of COVID-19 was categorized as mild, severe, or critically ill. Mild included non-pneumonia and mild pneumonia cases. Severe was characterized by dyspnea, respiratory frequency ≥30/min, blood oxygen saturation ≤93%, PaO_2_/FiO_2_ ratio <300, and/or lung infiltrates >50% within 24-48 hours. Critically ill cases were defined as respiratory failure requiring mechanic ventilation, septic shock, and/or multiple organ dysfunction/failure ^1,10^. The close date of the follow-up was April 1, 2020.

Patients with medical history of diabetes were defined by the diagnosis given by their doctors prior to the infection of SARS-CoV-2. A medical history of hypertension or blood pressure≥140/90mHg on admission were defined as any hypertension. The antidiabetic regimens were in principle unchanged during hospitalization whatever the patients had previous prescriptions before admission. Discontinuation or alteration of the antidiabetic treatment during hospitalization due to the clinical presentation of COVID-19 was at their physician’s discretion.

### Statistical Analysis

Continuous variables with normal distribution were expressed as mean (standard deviation) and compared using independent student’s t-test, and those with skewed distribution were expressed as median (interquartile range) and compared using Mann-Whitney U test. Categorical variables were presented as frequencies and percentages and compared using Fisher’s exact test. Survival time was estimated by the Kaplan-Meier method and Cox’s proportional hazard model was used to assess effects of diabetic status and covariates on mortality during hospitalization. Binary logistical regression analysis was used to assess the association between diabetic status and covariates and secondary endpoints, and the results were presented as odds ratios (ORs) with 95% confidence intervals (CIs). Only those variables that affected the hazard ratios (HRs) or ORs of endpoints with a *P* value <0.05 were included in the multivariate analysis. We excluded variables from the univariable analysis if the number of events was too small to calculate ORs or HRs. Restricted cubic splines were used to explore the relationship between blood glucose and hazard of death. As the relationship of predicted blood glucose was approximately linear above and below their cutoff, a linear spline model with a node at cutoff was fitted to calculate hazard ratios (HR) per mmol/L increase in blood glucose. Analyses were performed using SPSS version 25.0 software (SPSS, Inc, Chicago, Illinois, USA) or R-project (R Foundation, Vienna, Austria). A two-sided p value less than 0.05 was considered as statistically significant.

## RESULTS

In total, 2877 hospitalized patients with laboratory-confirmed COVID-19 were consecutively enrolled in the study. Baseline characteristics are presented in Table 1. Among these patients, 56/2877(1.9%) died in hospital and 387/2877(13.5%) had history of diabetes. Compared with patients with non-death, patients with death were older and more likely to be male, had higher prevalence of comorbidities such any hypertension on admission, diabetes, myocardial angina, previous PCI/CABG, and more often to have medications for diabetes and hypertension.

**Table 1.**
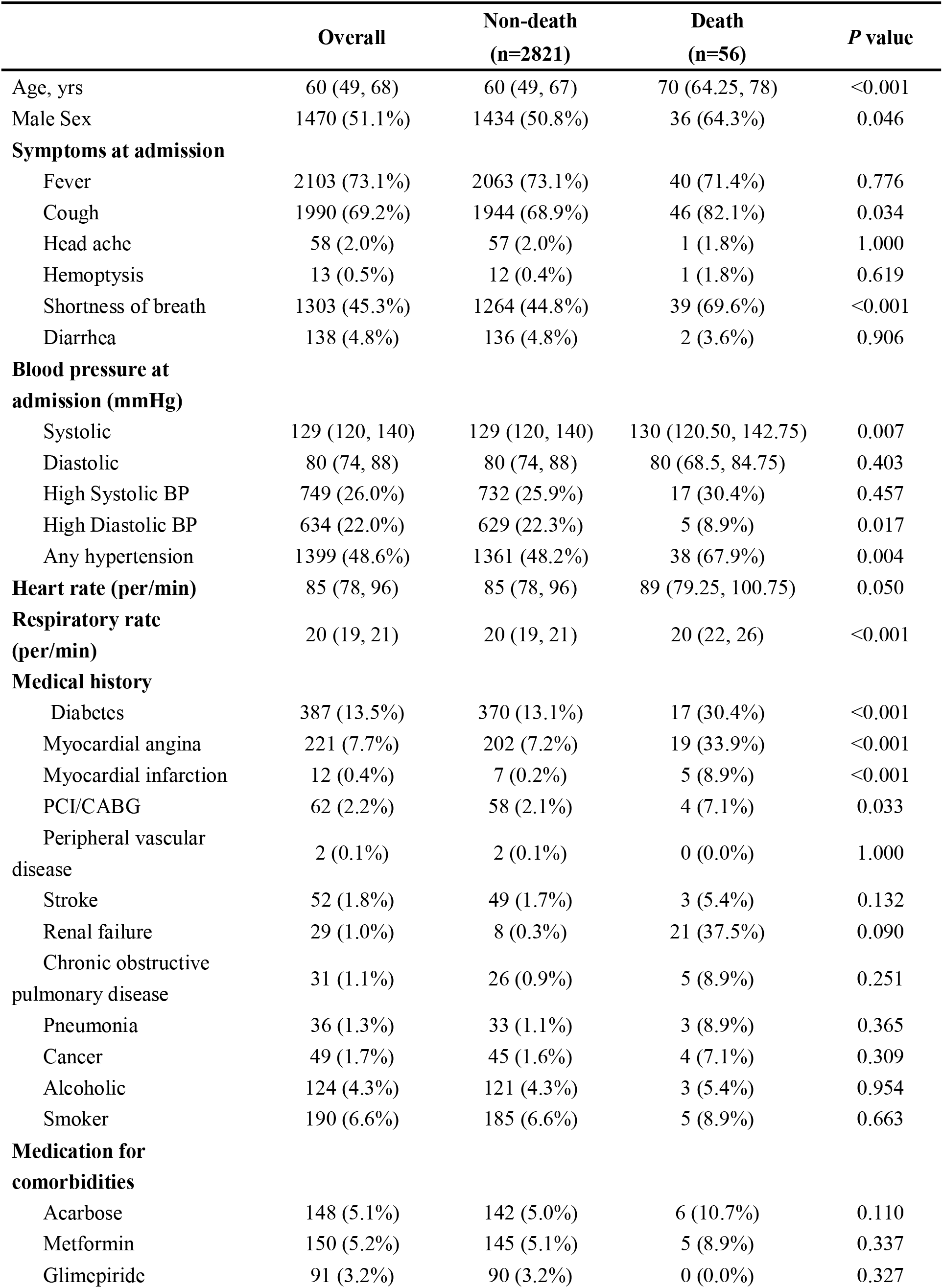
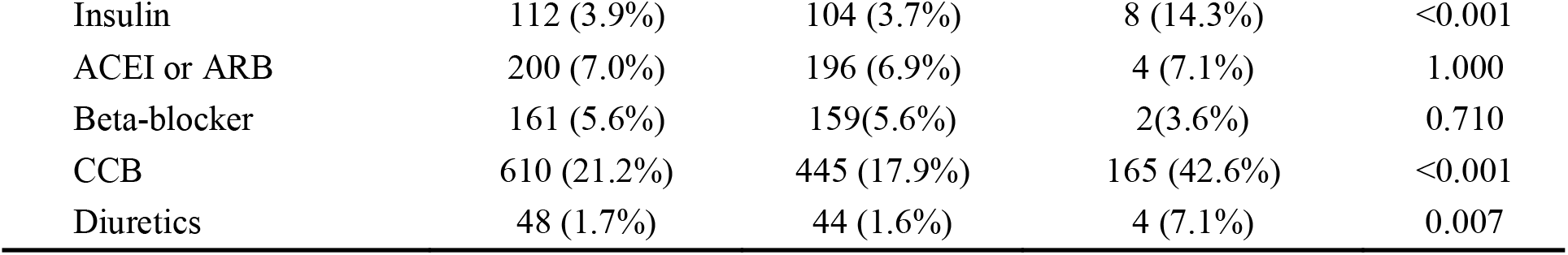
Baseline characteristics in patients with COVID-19.

|  | **Overall** | **Non-death**<br>**(n=2821)** | **Death**<br>**(n=56)** | **P value** |
| --- | --- | --- | --- | --- |
| Age, yrs | 60 (49, 68) | 60 (49, 67) | 70 (64.25, 78) | <0.001 |
| Male Sex | 1470 (51.1%) | 1434 (50.8%) | 36 (64.3%) | 0.046 |
| **Symptoms at admission** |  |  |  |  |
| Fever | 2103 (73.1%) | 2063 (73.1%) | 40 (71.4%) | 0.776 |
| Cough | 1990 (69.2%) | 1944 (68.9%) | 46 (82.1%) | 0.034 |
| Head ache | 58 (2.0%) | 57 (2.0%) | 1 (1.8%) | 1.000 |
| Hemoptysis | 13 (0.5%) | 12 (0.4%) | 1 (1.8%) | 0.619 |
| Shortness of breath | 1303 (45.3%) | 1264 (44.8%) | 39 (69.6%) | <0.001 |
| Diarrhea | 138 (4.8%) | 136 (4.8%) | 2 (3.6%) | 0.906 |
| **Blood pressure at admission (mmHg)** |  |  |  |  |
| Systolic | 129 (120, 140) | 129 (120, 140) | 130 (120.50, 142.75) | 0.007 |
| Diastolic | 80 (74, 88) | 80 (74, 88) | 80 (68.5, 84.75) | 0.403 |
| High Systolic BP | 749 (26.0%) | 732 (25.9%) | 17 (30.4%) | 0.457 |
| High Diastolic BP | 634 (22.0%) | 629 (22.3%) | 5 (8.9%) | 0.017 |
| Any hypertension | 1399 (48.6%) | 1361 (48.2%) | 38 (67.9%) | 0.004 |
| **Heart rate (per/min)** | 85 (78, 96) | 85 (78, 96) | 89 (79.25, 100.75) | 0.050 |
| **Respiratory rate (per/min)** | 20 (19, 21) | 20 (19, 21) | 20 (22, 26) | <0.001 |
| **Medical history** |  |  |  |  |
| Diabetes | 387 (13.5%) | 370 (13.1%) | 17 (30.4%) | <0.001 |
| Myocardial angina | 221 (7.7%) | 202 (7.2%) | 19 (33.9%) | <0.001 |
| Myocardial infarction | 12 (0.4%) | 7 (0.2%) | 5 (8.9%) | <0.001 |
| PCI/CABG | 62 (2.2%) | 58 (2.1%) | 4 (7.1%) | 0.033 |
| Peripheral vascular disease | 2 (0.1%) | 2 (0.1%) | 0 (0.0%) | 1.000 |
| Stroke | 52 (1.8%) | 49 (1.7%) | 3 (5.4%) | 0.132 |
| Renal failure | 29 (1.0%) | 8 (0.3%) | 21 (37.5%) | 0.090 |
| Chronic obstructive pulmonary disease | 31 (1.1%) | 26 (0.9%) | 5 (8.9%) | 0.251 |
| Pneumonia | 36 (1.3%) | 33 (1.1%) | 3 (8.9%) | 0.365 |
| Cancer | 49 (1.7%) | 45 (1.6%) | 4 (7.1%) | 0.309 |
| Alcoholic | 124 (4.3%) | 121 (4.3%) | 3 (5.4%) | 0.954 |
| Smoker | 190 (6.6%) | 185 (6.6%) | 5 (8.9%) | 0.663 |
| **Medication for comorbidities** |  |  |  |  |
| Acarbose | 148 (5.1%) | 142 (5.0%) | 6 (10.7%) | 0.110 |
| Metformin | 150 (5.2%) | 145 (5.1%) | 5 (8.9%) | 0.337 |
| Glimepiride | 91 (3.2%) | 90 (3.2%) | 0 (0.0%) | 0.327 |
| Insulin | 112 (3.9%) | 104 (3.7%) | 8 (14.3%) | <0.001 |
| ACEI or ARB | 200 (7.0%) | 196 (6.9%) | 4 (7.1%) | 1.000 |
| Beta-blocker | 161 (5.6%) | 159(5.6%) | 2(3.6%) | 0.710 |
| CCB | 610 (21.2%) | 445 (17.9%) | 165 (42.6%) | <0.001 |
| Diuretics | 48 (1.7%) | 44 (1.6%) | 4 (7.1%) | 0.007 |

Compared with patients with non-death, patients with death had higher respiratory rate and more likely presented with cough and shortness of breath. As shown in Figure 1 and Table 2, there are 17/387 (4.4%) died in the diabetic patients and 39/2490 (1.6%) died in non-diabetic patients (*P*<0.001). After adjusted for confounders, diabetic patients were still associated with a 2-fold increase in the hazard of death as compared to non-diabetic patients (adjusted HR 2.11, 95%CI: 1.16-3.83, *P*=0.014). The baseline characteristics of patients with or without secondary endpoints (the severity of COVID-19 and use of invasive mechanical ventilation) were presented in supplemental Table 1. As compared with patients with mild COVID-19, the patients with severe/critical COVID-19 had higher prevalence of diabetes (144/744, 19.4% vs 243/2133,11.4%, *P*<0.001). After adjusted for confounders, diabetes was still associated with an increase in the odds of developing a severe/critical COVID-19 as compared to non-diabetic patients (adjusted OR1.48, 95%CI: 1.17-1.87, *P*=0.001) (supplemental Table 2).As compared with patients without receiving invasive mechanical ventilation, the patients receiving invasive mechanical ventilation had higher prevalence of diabetes (17/65, 26.2% vs 370/2812, 13.2% *P*=0.002)(supplemental Table1). After adjusted for confounders, diabetes was not associated with an increase in the odds of receiving invasive mechanical ventilation as compared to non-diabetic patients (supplemental table 3).

**Table 2.** Association between diabetes and death in patients with COVID-19.

|  | Univariable |  | Multivariable |  |
| --- | --- | --- | --- | --- |
|  | HR (95%CI) | *P* value | HR (95%CI) | *P* value |
| Diabetes | 2.70 (1.53, 4.77) | 0.001 | 2.11 (1.16, 3.83) | 0.014 |
| Age (year) | 1.08 (1.05, 1.10) | <0.001 | 1.08 (1.05, 1.11) | <0.001 |
| Sex (male) | 1.78 (1.03, 3.07) | 0.039 |  |  |
| Any hypertension | 2.21 (1.26, 3.87) | 0.006 |  |  |
| Myocardial angina | 6.49 (3.73, 11.30) | <0.001 | 3.29 (1.81, 6.00) | <0.001 |
| Previous PCI/CABG* | 4.20 (1.52, 11.64) | 0.006 |  |  |
| Cough | 1.88 (0.95, 3.73) | 0.071 | 2.54 (1.27, 5.08) | 0.008 |
| Shortness of breath | 2.54 (1.43, 4.49) | 0.001 |  |  |
| Respiratory rate (per/min) | 1.78 (1.03, 3.07) | 0.039 | 1.28 (1.22, 1.35) | <0.001 |

\* PCI=Percutaneous Coronary Intervention; CABG= Coronary Artery Bypass Grafting

**Table 3.**
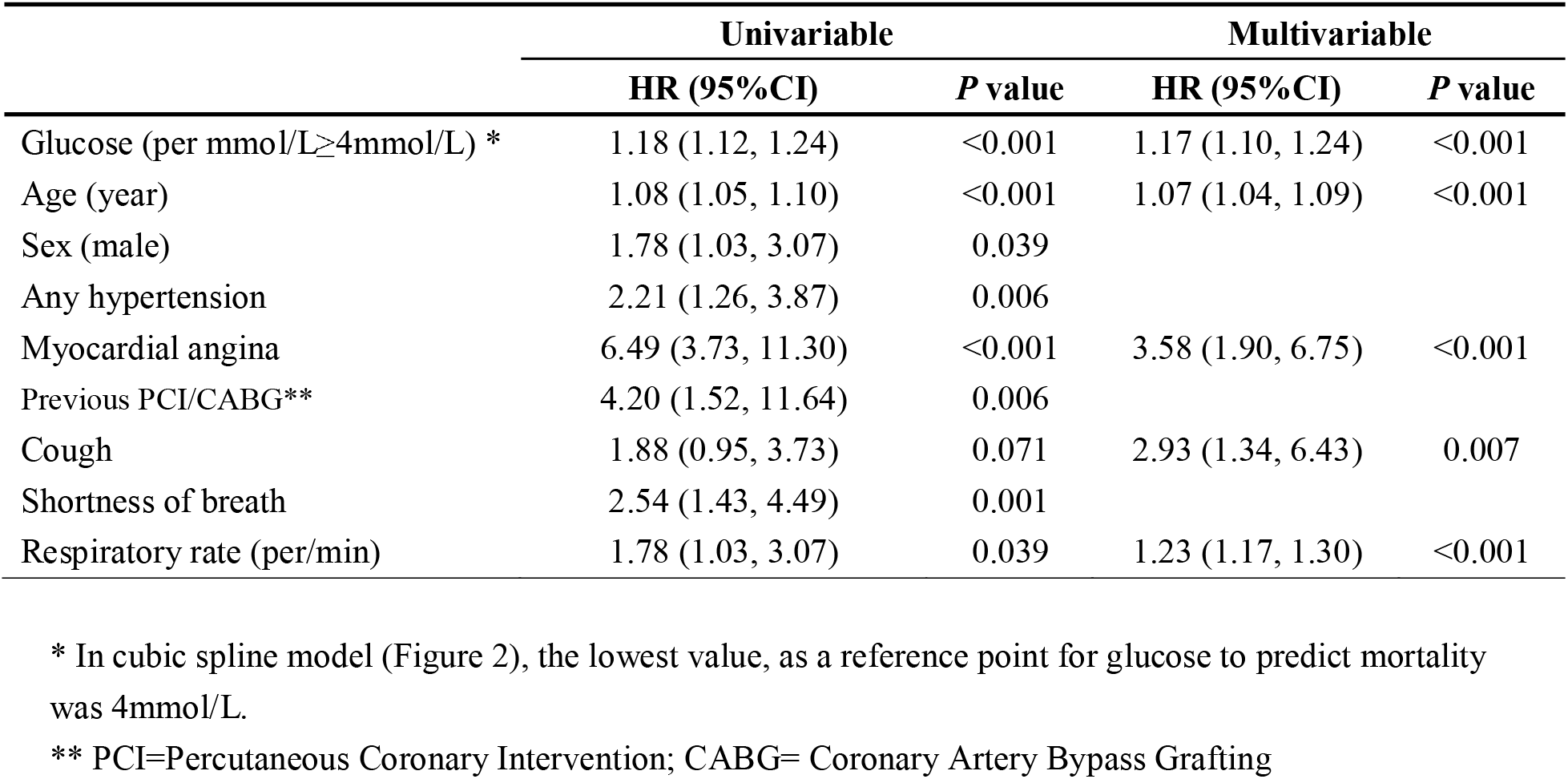
Association between glucose and mortality in COVID-19 patients.

Restricted cubic splines were used to explore the possible relationship between blood glucose and mortality. As shown in figure 2, U-shaped association between blood glucose and mortality was observed in the overall population cohort, and the lowest value, as a reference point for glucose to predict mortality was 4mmol/L. As shown in table 3, when glucose was more than 4mmol/L, it was significantly associated with subsequent mortality on COVID-19 (adjusted HR 1.17, 95%CI: 1.10-1.24, *P*=0.002).

**Figure 1.**
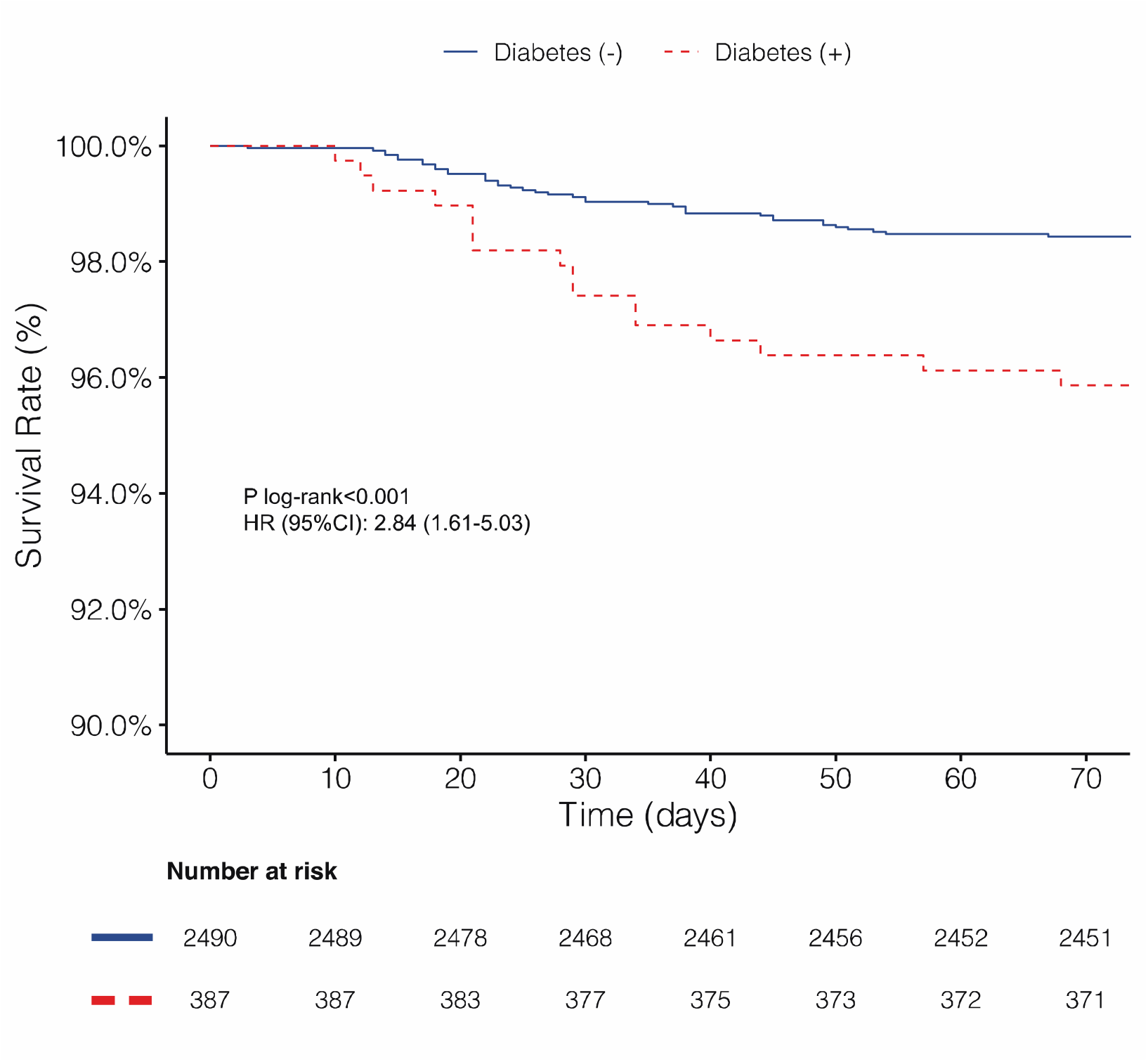
Kaplan-Meier survival curves for mortality in patients with or without diabetes.

**Figure 2.**
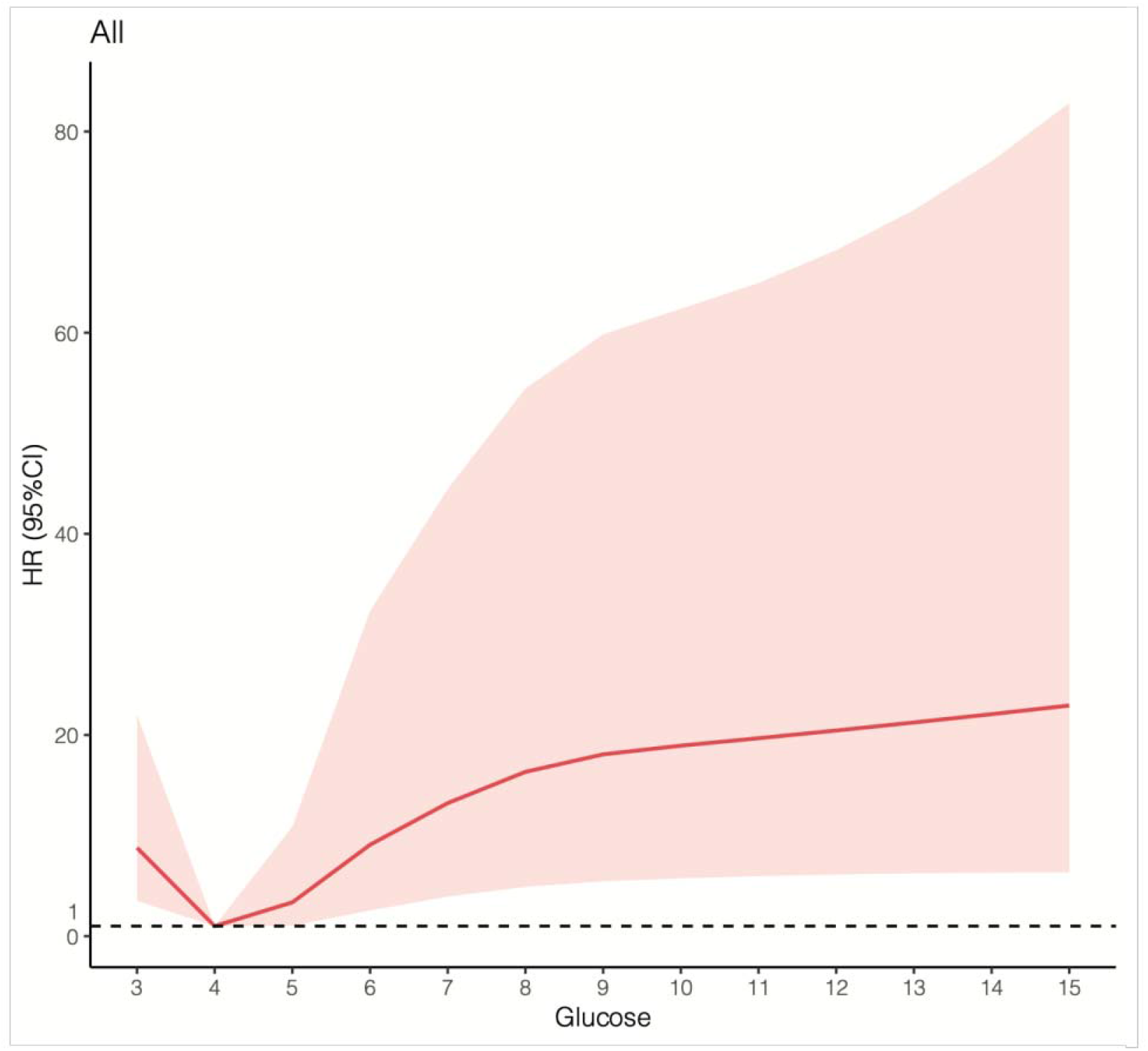
Association between glucose and hazard ratio of mortality in an adjusted cubic spline in patients with COVID-19.

Among the 387 patients with diabetes, there were 102/387 (26.4%) diabetic patients treated with insulin, 140/387 (36.2%) with acarbose and 142/387 (36.7%) with metformin. No statistically significant differences were found in the mortality between the Insulin and non-insulin treated patients (8/102, 7.8% vs 9/285, 3.2% *P*=0.089), metformin and non-metformin treated patients (5/142, 3.5% vs 12/245, 4.9% *P*=0.524), and acarbose and non-acarbose treated patients (6/140, 4.3% vs 11/247, 4.5% *P*=0.938).

## DISCUSSION

In the presented analysis, we investigated the association of the diabetes and blood glucose on mortality in patients with COVID-19. The salient findings from the present study are summarized as follows:

1. After adjustment for the confounders, compared with non-diabetic patients, the diabetic patients were still associated with a 2-fold increase in mortality.
2. The on-admission blood glucose (per mmol/L≥4mmol/L) was significantly associated with subsequent mortality on COVID-19.

Previous reports have suggested that diabetes is one of the most common comorbidities of the COVID-19 patients and might have influenced the prognosis^11-13^. The estimated incidence of diabetes in the COVID-19 patients ranged from 9.7% to 20%^11-13^. COVID-19 patients with diabetes may have poor prognosis, evidenced by the fact that the incidence of diabetes was two-fold higher in ICU/severe cases than in their non-ICU/severe counterparts^13^. Although the evidence is still limited, studies have suggested that diabetes had a hazard ratio of 2-3 for mortality^11-13^. However, none of these previous studies have adjusted for confounding factors, such as age, hypertension, or cardio-cerebrovascular diseases, which have been previously identified as predictors of COVID-19 related death. Recently, a study including 191 COVID-19 found that patients with diabetes has a hazard ratio of 2.85 for death^2^. However, after adjusting for potential confounding factors, the corresponding risk disappeared. The small sample size maybe the main reason for the neutral results. In the present study, with a large-scale cohort of 2877 patients with COVID-19, we demonstrated that diabetes was associated with a high risk of mortality during hospitalization after adjusting for the underlying confounders. To our knowledge, this is the first study to report diabetes as an independent risk factor for mortality in patients with COVID-19.

We found that the proportion of on-admission glucose (fasting glucose) above 7mmol/L, as one of a criteria to diagnose diabetes, was 13.5% ^14^. Although possibility that inadequate anti-medications were prescribed to the diabetic patients or that undiagnosed diabetes were present, we still speculated that the dysglycemia was frequent in patient with COVID-19 given the large proportion of patients with abnormal glucose value.

Large body of literature has found that dysglycemia including hyperglycemia, hypoglycemia and the increased glucose variability is independently associated with mortality in critically ill patients^15,16^.Furthermore, there were evidences showing that the blood glucose level on admission is also independently associated with adverse outcomes in patients with community acquired pneumonia (CAP). Earlier, admission glucose level >14 mmol/l was identified as one of the 20 factors associated with poor outcomes in CAP^6^. Later on, a prospective cohort study conducted by Finlay A et al. found that, even after adjustment for confounding factors, a lower glucose level >11 mmol/l was also remained significantly associated with subsequent adverse outcomes in patients with CAP^5^. In 2009, when the pandemic influenza A (H1N1) virus spread worldwide, wang et al. demonstrated that the fasting blood glucose was an independent predictor for severity of H1N1 pneumonia^7^. It has been inferred that the reasons for the hyperglycemia in H1N1-mediated pneumonia are as follows: 1. By a direct pathway, H1N1 virus could potentially damage pancreatic β-cells evidenced by downregulated homa-β in the H1N1 infected patients^7^; 2. By an indirect pathway, H1N1-mediated elevations of FPG might be the result of multiple factors’ combinations caused by virus infection, such as inflammatory stress, multiple organ impairment, and so on, while the changes of plasma glucose were not caused by H1N1 actions on insulin secretion by pancreatic β-cells or decreasing sensitivity to insulin^7^.

Although there is no direct evidence that pancreatic β-cells is also a target for SARS-CoV-2, the dysglycemia is not uncommon in patients with COVID-19. However, whether the dysglycemia is a predictor for clinical outcomes in patients with COVID-19 remains unknown. In the present analysis, we used restricted cubic splines and Cox regression to investigate the predictive role of dysglycemia for clinical outcomes. We found that after adjusted for the cofounders, the admission blood glucose (per mmol/L≥4mmol/L) was significantly associated with subsequent mortality on COVID-19. For each 1-mmol/l increase in admission glucose, risk of in-hospital mortality increases 17% in patients with COVID-19. Therefore, a more active intervention should be taken to control the blood glucose. Additionally, the hyperglycemia on admission maybe a strong stratification factor to differentiate high risk patients with COVID-19.

According to our present data, the glucose control should be emphasized. Unfortunately, few studies have been conducted to investigate the impact of anti-diabetic medications on the mortality of COVID-19. So far, no specific anti-viral treatment exists, hence, a range of existing host-directed therapies that have proven to be safe could potentially be repurposed to treat 2019-nCoV infection^17^. Several marketed drugs with safety profiles such as metformin, glitazones could reduce immunopathology, boost immune responses, and prevent or curb ARDS^18^. In the present study, we retrospectively investigate the effect of anti-diabetic medications including metformin regarded as a medication for host-directed therapies on the mortality in patients with COVID-19. No significant difference was observed as to the mortality and other clinical endpoints in the COVID-19 patients with or without the above medications.

However, the analysis is underpowered, which might lead to the neutral results. Future dedicated study with adequate sample size is needed to further investigate such issue.

## Limitations

First, since our results were based only on admission glucose, it was likely that we had underestimated the risks associated with hyperglycemia given that the patients with the highest levels of glucose would have been more likely to be treated in the hospital setting.

Second, we did not routinely collect measures of long-term glucose control, such as HbA1c, which would have allowed us to distinguish long-term poorly controlled diabetic patients from those with hyperglycemia as a measure of significant physiological stress.

Third, even though we analyzed one of the largest cohorts of hospitalized patients with COVID-19 up to date, we had insufficient numbers to examine the role of various anti-glycemic treatments (e.g., insulin, metformin) on outcomes. Our findings should be interpreted cautiously. Larger scale and dedicated cohort study or RCT were needed to verify our conclusions.

## CONCLUSIONS

In conclusion, diabetes was independently associated with increased mortality, and on-admission blood glucose (per mmol/L≥4mmol/L) was also an independent factor for mortality in patients with COVID-19. The mortality was not affected by the anti-diabetic medications. These data support that, regardless of anti-diabetic medications administrated, proper blood glucose control should be achieved for possibly better outcomes in patients with COVID-19. However, due to the observational nature of the study, the results should be interpreted cautiously.

## Data Availability

We would like to provide all data referred to the manuscript if they are requested.

