## Supplemental table 1 for "Effects of Diabetes and Blood Glucose on COVID-19 Mortality: A Retrospective Observational Study"

**Supplemental table 1. Baseline characteristics in COVID-19 patients with or without secondary endpoint.**

|  | **Overall** | **Pneumonia type**  **(Mild)** | **Pneumonia type**  **(Severe/critical)** | ***P* value** | **Non-** **invasive mechanical ventilation** | **Invasive mechanical ventilation** | ***P* value** |
| --- | --- | --- | --- | --- | --- | --- | --- |
|  |  | **(n=2133)** | **(n=744)** |  | **(n=2812)** | **(n=65)** |  |
| Age, yrs | 60 (49, 68) | 58 (47, 66) | 64 (55, 72) | <0.001 | 59.5 (49, 67) | 69 (63.5, 75.5) | <0.001 |
| Male Sex | 1470 (51.1%) | 1070 (50.2%) | 400 (53.8%) | 0.091 | 1426 (50.7%) | 44 (67.7%) | 0.007 |
| **Symptoms at admission** |  |  |  |  |  |  |  |
| Fever | 2103 (73.1%) | 1531 (71.8%) | 572 (76.9%) | 0.007 | 2057 (73.2%) | 46 (70.8%) | 0.669 |
| Cough | 1990 (69.2%) | 1444 (67.7%) | 546 (73.4%) | 0.004 | 1938 (68.9%) | 52 (80.0%) | 0.056 |
| Head ache | 58 (2.0%) | 45 (2.1%) | 13 (1.7%) | 0.545 | 58 (2.0%) | 1 (1.5%) | 1.000 |
| Hemoptysis | 13 (0.5%) | 8 (0.4%) | 5 (0.7%) | 0.470 | 12 (0.4%) | 1 (1.5%) | 0.700 |
| Shortness of breath | 1303 (45.3%) | 904 (42.4%) | 399 (53.6%) | <0.001 | 1258 (44.7%) | 45 (69.2%) | <0.001 |
| Chest pain | 57 (2.0%) | 41 (1.9%) | 16 (2.2%) | 0.700 | 49 (1.7%) | 8 (12.3%) | <0.001 |
| Diarrhea | 138 (4.8%) | 104 (4.9%) | 34 (4.6%) | 0.737 | 136 (4.8%) | 2 (3.1%) | 0.717 |
| Shivering | 51 (1.8%) | 35 (1.6%) | 16 (2.2%) | 0.364 | 44 (1.6%) | 7 (10.8%) | <0.001 |
| **Blood pressure at admission (mmHg)** |  |  |  |  |  |  |  |
| Systolic | 129 (120, 140) | 128 (120, 139) | 130 (120, 143) | <0.001 | 129 (120, 140) | 130 (120,143.5) | 0.208 |
| Diastolic | 80 (74, 88) | 80 (74, 88) | 80 (74, 88) | 0.827 | 80 (74, 88) | 80 (68 88) | 0.037 |
| High Systolic BP | 749 (26.0%) | 506 (23.7%) | 243 (32.7%) | <0.001 | 728 (25.9%) | 21 (32.3%) | 0.244 |
| High Diastolic BP | 634 (22.0%) | 469 (21.8%) | 165 (23.5%) | 0.914 | 626 (22.3%) | 8 (12.3%) | 0.056 |
| Hypertensive | 1399 (48.6%) | 969 (45.1%) | 430 (71.6%) | <0.001 | 1355 (48.2%) | 44 (67.7%) | 0.002 |
| **Heart rate (per/min)** | 85 (78, 96) | 84 (78, 95) | 86 (78, 98) | 0.007 | 85 (78, 96) | 90 (80.5, 101) | 0.003 |
| **Respiratory rate (per/min)** | 20 (19, 21) | 20 (19, 21) | 20 (20, 22) | <0.001 | 20 (19, 21) | 22 (20, 25) | <0.001 |
| **Medical history** |  |  |  |  |  |  |  |
| Diabetes | 387 (13.5%) | 243 (11.4%) | 144 (19.4%) | <0.001 | 370 (13.2%) | 17 (26.2%) | 0.002 |
| Myocardial angina | 221 (7.7%) | 131 (6.1%) | 90 (12.1%) | <0.001 | 204 (7.3%) | 17 (26.2%) | <0.001 |
| Myocardial infarction | 12 (0.4%) | 8 (0.4%) | 4 (0.5%) | 0.793 | 7 (0.2%) | 5 (7.7%) | <0.001 |
| PCI/CABG | 62 (2.2%) | 35 (1.6%) | 27 (3.6%) | 0.001 | 60 (2.1%) | 2 (3.1%) | 0.932 |
| Peripheral vascular disease | 2 (0.1%) | 1 (0.0%) | 1 (0.1%) | 0.450 | 2 (0.1%) | 0 (0%) | 1.000 |
| Stroke | 52 (1.8%) | 34 (1.6%) | 18 (2.4%) | 0.143 | 49 (1.7%) | 3 (4.6%) | 0.212 |
| Renal failure | 29 (1.0%) | 5 (0.2%) | 24 (3.2%) | <0.001 | 6 (0.2%) | 23 (35.4%) | <0.001 |
| Chronic obstructive pulmonary disease | 31 (1.1%) | 13 (0.6%) | 18 (2.4%) | <0.001 | 24 (0.9%) | 7 (10.8%) | <0.001 |
| Pneumonia | 36 (1.3%) | 15 (0.7%) | 21 (2.8%) | <0.001 | 29 (0%) | 7 (10.8%) | <0.001 |
| Cancer | 49 (1.7%) | 34 (1.6%) | 15 (2.0%) | 0.444 | 44 (1.6%) | 5 (7.7%) | <0.001 |
| Alcoholic | 124 (4.3%) | 96 (4.5%) | 28 (3.8%) | 0.394 | 120 (4.3%) | 4 (6.2%) | 0.666 |
| Smoker | 190 (6.6%) | 140 (6.6%) | 50 (6.7%) | 0.882 | 184 (6.5%) | 6 (9.2%) | 0.542 |
| **Medication for comorbidities** |  |  |  |  |  |  |  |
| Acarbose | 148 (5.1%) | 98 (4.6%) | 50 (6.7%) | 0.024 | 140 (5.0%) | 8 (12.3%) | 0.018 |
| Metformin | 150 (5.2%) | 102 (4.8%) | 48 (6.5%) | 0.078 | 144 (5.1%) | 6 (9.2%) | 0.233 |
| Glimepiride | 91 (3.2%) | 62 (2.9%) | 29 (3.9%) | 0.183 | 91 (3.2%) | 0 (0%) | 0.265 |
| Insulin | 112 (3.9%) | 64 (3.0%) | 48 (6.5%) | <0.001 | 107 (3.8%) | 5 (7.7%) | 0.201 |
| ACEI or ARB | 200 (7.0%) | 121 (5.7%) | 79 (10.6%) | <0.001 | 195 (6.9%) | 5 (7.7%) | 1.000 |
| Beta-blocker | 161 (5.6%) | 108 (5.1%) | 53 (7.1%) | 0.035 | 158 (5.6%) | 3 (4.6%) | 0.940 |
| CCB | 610 (21.2%) | 397 (18.6%) | 213 (28.6%) | <0.001 | 582 (20.7%) | 28 (43.1%) | <0.001 |
| Diuretics | 48 (1.7%) | 27 (1.3%) | 21 (2.8%) | 0.004 | 44 (1.6%) | 4 (6.2%) | 0.018 |
