## Supplemental table 2 for "Effects of Diabetes and Blood Glucose on COVID-19 Mortality: A Retrospective Observational Study"

**Supplemental table 2. Association between diabetes and pneumonia type in patients with COVID-19**

|  | **Univariable** | | **Multivariable** | |
| --- | --- | --- | --- | --- |
|  | **HR (95%CI)** | ***P* value** | **HR (95%CI)** | ***P* value** |
| Diabetes | 1.87 (1.49, 2.34) | <0.001 | 1.48 (1.17, 1.87) | 0.001 |
| Age | 1.03 (1.03, 1.04) | <0.001 | 1.03 (1.04, 1.03) | <0.001 |
| Hypertension | 1.65 (1.39, 1.95) | <0.001 | 1.29 (1.08, 1.54) | 0.015 |
| Myocardial angina | 2.10 (1.59, 2.79) | <0.001 | 1.44 (1.07, 1.94) | 0.015 |
| Previous PCI/CABG | 2.26 (1.36, .76) | 0.002 |  |  |
