## Supplemental table 3 for "Effects of Diabetes and Blood Glucose on COVID-19 Mortality: A Retrospective Observational Study"

**Supplemental table 3. Association between diabetes and invasive mechanical ventilation in patients with COVID-19**

|  | **Univariable** | | **Multivariable** | |
| --- | --- | --- | --- | --- |
|  | **HR (95%CI)** | ***P* value** | **HR (95%CI)** | ***P* value** |
| Diabetes | 2.34 (1.33, 4.11) | 0.003 |  |  |
| Age | 1.07 (1.05, 1.09) | <0.001 | 1.06 (1.04, 1.08) | <0.001 |
| Hypertension | 2.25 (1.33, 3.81) | 0.002 |  |  |
| Myocardial angina | 4.53 (2.56, 8.02) | <0.001 | 2.73 (1.51, 4.94) | 0.001 |
